# Heated tobacco products and a telemedicine smoking cessation programme: A retrospective study

**DOI:** 10.1101/2022.02.15.22270564

**Authors:** Akihiro Nomura, Takaaki Ikeda, Toshiki Fujimoto, Yusaku Morita, Chie Taniguchi, Tetsuro Ishizawa, Takahiro Tabuchi

## Abstract

**Objectives:** Japan is one of the largest markets for heated tobacco products (HTP), and the number of HTP users, including dual users, is burgeoning. However, it is not yet clear whether a telemedicine smoking cessation programme is helpful for nicotine-dependent people who use HTPs.

**Methods:** We retrospectively evaluated the effectiveness of a telemedicine smoking cessation programme for nicotine-dependent tobacco product users, comparing short- and long-term continuous abstinence rates (CAR) from 9 to 24 weeks (CAR9-24) and 9 to 52 weeks (CAR9-52). We divided programme participants into 1) exclusively cigarette users, 2) exclusively HTP users, and 3) dual users. Using logistic regression with inverse probability weighting, an adjusted odds ratio (aOR) with a 95% confidence interval (CI) for CAR was calculated to compare the differences among the three groups.

**Results:** We analysed 733 telemedicine smoking cessation programme participants (exclusively cigarette users, 52%; exclusively HTP users, 31%; and dual-users, 16%) dating August 2018 to October 2020. HTP users had higher CARs than the exclusively cigarettes group in CAR9-24 (aOR, 1.12; CI, 1.02–1.23; p=0.02) and CAR9-52 (aOR 1.09; CI, 0.99–1.19; p=0.08). Conversely, dual users had lower CARs than the exclusively cigarettes group in the CAR9-24 (aOR, 0.85; CI, 0.76–0.95; p=0.004) and CAR9-52 (aOR, 0.88; CI, 0.79–0.97; p=0.01).

**Conclusions:** Exclusively HTP users achieved higher CARs, whereas dual users had lower CARs than exclusively cigarette users over short- and long-term periods. A telemedicine smoking cessation programme may be a reasonable option for exclusively HTP users.

**Key messages:** *What is already known on this topic:* Telemedicine smoking cessation programmes can be helpful for some nicotine-dependent people.

*What this study adds:* Exclusively HTP users had higher continuous abstinence rates (CARs) than the exclusively cigarettes group. Conversely, dual users had lower CARs than the exclusively cigarettes group. A telemedicine smoking cessation programme may be a reasonable option for exclusive HTP users.

*How this study might affect research, practice, or policy:* In addition to conventional combustible cigarettes, the number of HTP users, including dual users, is burgeoning in Japan. This study could add an effective and alternative option for HTP users to remotely quit smoking in the era of COVID-19 and beyond.

## INTRODUCTION

Smoking is a major preventable risk factor for cardiovascular diseases, respiratory diseases, and malignant neoplasms^1,2^. Additionally, it is the leading cause of death from non-communicable diseases and external causes in Japan and other countries. In addition to conventional cigarette smoking, the number of heated tobacco products (HTP) users is increasing, especially in Japan, since 2014 when Philip Morris International first released a HTP, IQOS, in Japan and Italy^3^. HTPs are electronic devices that heat tobacco leaves instead of combusting them to produce aerosols^4^. One serious problem with HTP usage is that the exclusive or dual use of heated or e-cigarettes is misconstrued as an alternative to smoking cessation treatment^5^, which can lead to nicotine dependence in cigarette smokers and hinder the promotion of smoking cessation^6^. Since more than 20 million smokers in Japan use conventional cigarettes, HTP, or dual products^3,7^, and there are up to 130,000 smoking-related deaths per year^8^, smoking cessation is expected to significantly reduce the occurrence of the various diseases mentioned above, extend life expectancy, and ultimately reduce mortality rates and medical costs^8,9^.

In Japan, nicotine-dependent patients, including HTP users, are eligible for a 12-week standard smoking cessation treatment programme with clinic visits and an initial screening interview in which their exhaled carbon monoxide levels are checked^10^. Moreover, some commercial telemedicine smoking cessation programmes are available for members of corporate health insurance societies (a public body that provides health insurance to corporation employees)^11^. However, biochemically check smoking cessation status through exhaled carbon monoxide, which is currently measured at every visit in a standard smoking cessation programme, is not effective for HTP use^12^. Moreover, it has not yet been assessed whether these smoking cessation programmes are helpful for smokers who use HTPs since they were initially made for cigarette smokers^10,11^.

Here, we tested the effectiveness of a telemedicine smoking cessation programme among nicotine-dependent patients who had used 1) exclusively conventional cigarettes; 2) exclusively HTPs; or 3) dual cigarettes and HTP products.

## METHODS

### Study overview

We conducted a retrospective study to assess the effectiveness of the telemedicine smoking cessation programme provided by Linkage Inc., Tokyo, Japan^13^. Among participants in the programme, we compared the success rates of smoking cessation between smokers who had used conventional cigarettes and those who had used HTPs. This study was performed in compliance with the Declaration of Helsinki, Ethical Guidelines for Medical and Health Research Involving Human Subjects, and all other applicable laws and guidelines in Japan. The study protocol was approved by the Kanazawa University institutional review board (2020-206 [3573]).

### Participants

The present study was conducted between August 2018 and October 2020 and enrolled Japanese current smokers aged 20 years or older who applied for the telemedicine smoking cessation programme through the health insurance association. All the participants previously answered self-administered questionnaires on smoking status including their smoking period, cigarette and/or HTP use, smoking cessation history, reasons for this smoking cessation programme (e.g., health consciousness, family or friend’s recommendations, financial burden, workplace smoking cessation policy, and convenience of telemedicine), medical complications and treatments, marital states, job type, and drinking habits. The participants were divided into three groups as 1) exclusively cigarette users, 2) exclusively HTP users, and 3) both cigarette and HTP dual users. We excluded smokers who had severe mental illness or severe physical comorbidities which required hospitalization during the program and those unavailable for the follow-up surveys.

### Outcomes

The primary outcomes were the participants’ continuous abstinence rates (CARs) from week 9 to 24 (CAR9-24) and week 9 to 52 (CAR9-52). The secondary outcomes were point abstinences that were defined as self-reported continuous abstinence from both cigarettes and HTP smoking since latest follow-up survey at week 12 (equal to CAR from week 9 to 12 [CAR9-12]), 24, 36, and 52. Smoking cessation success was confirmed with the structural interview performed by the telemedicine doctors at week 8 session and participant’s self-reports to the follow-up survey at weeks 12, 24, 36, and 52. CAR was defined as one or more consecutive weeks of abstinence since the telemedicine session finished at week 8. For example, if one participant answered that they had point abstinence at both weeks 12 and 24 via the follow-up survey, we considered that the participants achieved continuous abstinence from week 9 to 24 (CAR9-24). Participants who self-reported ‘relapse’ or did not respond to the follow-up survey (‘no reply’) were considered to have failed smoking cessation.

### The Linkage telemedicine smoking cessation programme

The Linkage telemedicine smoking cessation programme is a 52-week (12-month) completely remote online programme through the *D-CUBE* telemedicine app made by Linkage Inc. (Tokyo, Japan) (Figure 1). This programme consists of the 8-week (2-month) telemedicine programme provided by primary doctors and a 10-month follow-up period sending surveys and smoking cessation advice via the app at weeks 12, 24, 36, and 52. For the first eight weeks, four telemedicine sessions were conducted in-person through a videoconferencing system in the app, during which doctors evaluated participant’s physical and mental conditions, encouraged smoking cessation, and offered advice on quitting smoking. Each telemedicine treatment time was about 15 minutes. Smoking cessation medications, such as varenicline or nicotine patches, were also prescribed by doctors’ discretion considering participants’ baseline characteristics and were directly sent to the participant’s home or office place. After completing the 8-week telemedicine programme, the *D-CUBE* app sent follow-up surveys and smoking cessation advice at weeks 12, 24, 36, and 52. Smoking cessation specialists created the smoking cessation encouragement and facilitation advice messages.

**Figure 1.**
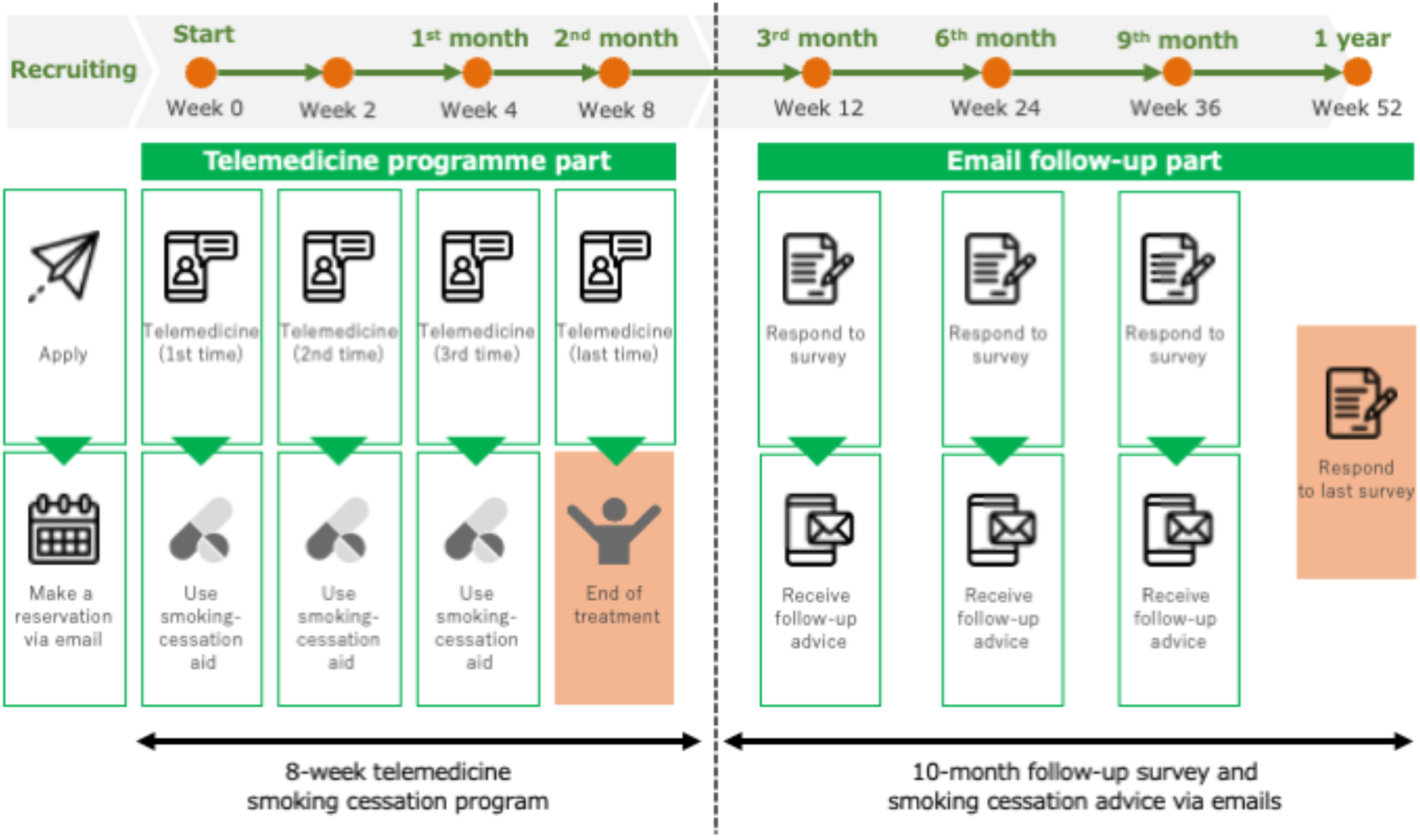
Overview of the Linkage telemedicine smoking cessation programme and the D-CUBE telemedicine app for smoking cessation.

### Data collection

We collected baseline information and follow-up surveys on each participant through the telemedicine app, including their age, sex, years spent smoking, number of cigarettes per day, and tobacco dependence screening score using the Tobacco Dependence Screener (TDS)^14^.

### Statistical Analysis

Continuous variables are presented as mean ± standard deviation or median (interquartile range [IQR]) depending on distribution. Categorical variables are presented as numbers and percentages. A random forest imputation algorithm was used to deal with missing variables^15^. For datasets with a mixture of categorical and continuous variables, random forest imputation enables imputation with less bias than other imputation methods, such as multiple imputations^15^. We used all survey variables not missing more than 30% of values in each variable for the imputation. Random forest imputation was conducted using Python version 3.8.3 with the *MissForest* package. In addition, we estimated inverse probability weights using generalised boosted models with 5,000 regression trees^16^. This model used the following parameters: age, sex, body mass index, years of smoking, past attempt for cessation, TDS score, regular alcohol intake, hypertension, diabetes mellitus, cardiovascular disease, mental disorders, varenicline use, and nicotine patch use. We showed the standardised mean differences for each parameter to assess the balance of the three tobacco product groups. A maximum standardised difference of more than 0.2 indicated an imbalance among groups. After balancing the covariates among the tobacco product groups, we tested the differences in outcomes using logistic regression. Analyses for the inverse probability weighting method were conducted using *R* software version 4.1.2 (R Foundation for Statistical Computing, Vienna, Austria) with a *twang* package^16^.

## RESULTS

We retrospectively analysed 750 participants of the Linkage telemedicine smoking cessation programme between August 2018 and October 2020. Of these, 17 participants were excluded due to unavailable smoking product usage data. Thus, we included 733 participants for further analyses (Figure 2). Table 1 shows the baseline characteristics of the participants before inverse probability weighting. The mean age was 42.4 ± 9.8 years, 94% were men, and the median years spent smoking was 22. The types of smoking products included exclusively combustible cigarettes (52%), exclusively HTPs (31%), and dual use of combustible cigarettes and HTPs (16%). In terms of smoking cessation medication, varenicline was prescribed to 90% of programme participants. Compared with exclusively cigarette users, exclusively HTP users were younger, had smoked for fewer years, more attempts for smoking cessation, and were prescribed varenicline more frequently than nicotine patches as smoking cessation medication. Additionally, dual users were younger and had smoked for fewer years compared to cigarette users. After inverse probability weighting, the maximum standardised differences among the tobacco product groups were all ≤0.15, indicating that the parameters were well balanced among the groups.

**Figure 2.**
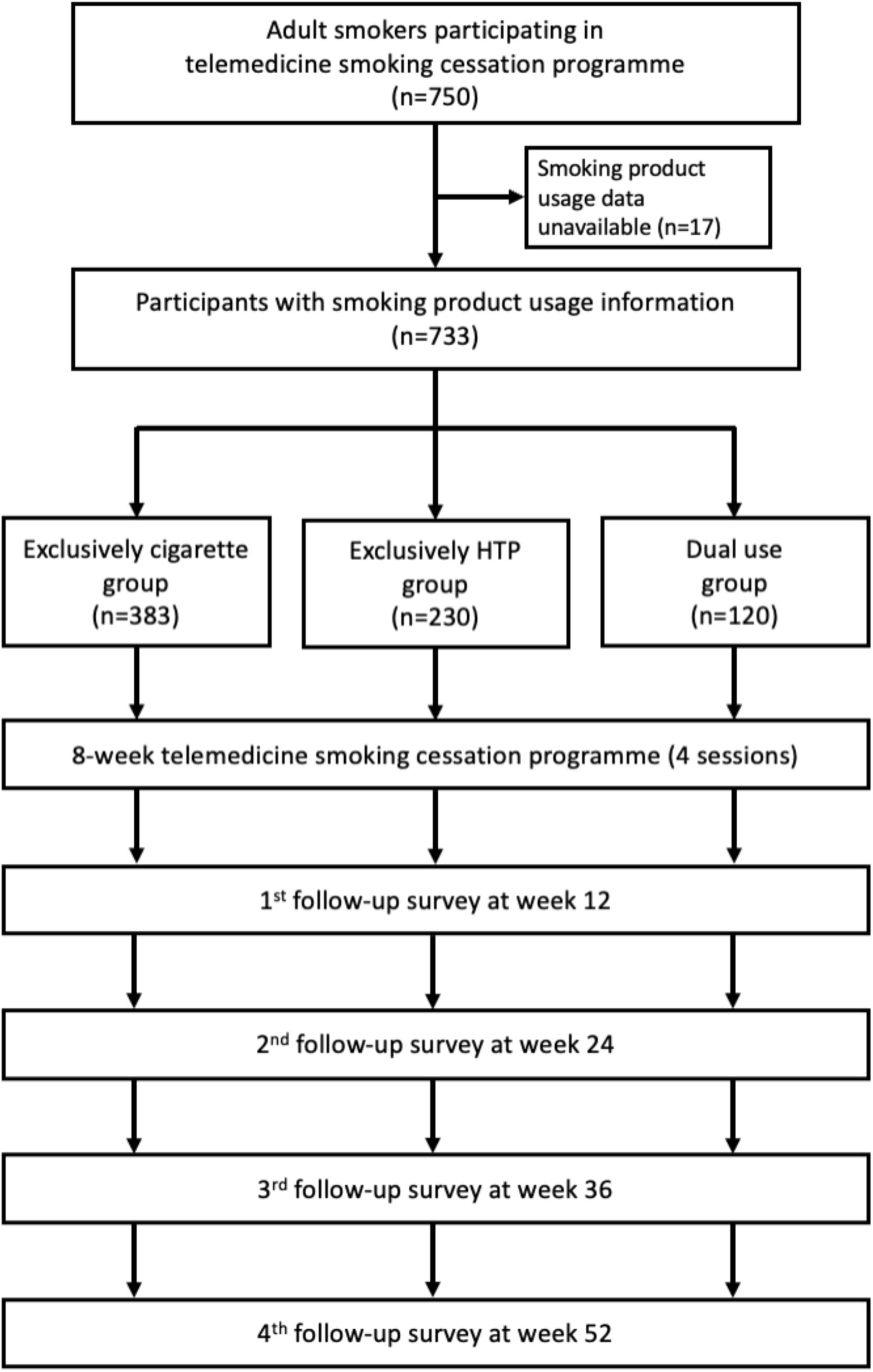
Study flowchart.

**Table 1.**
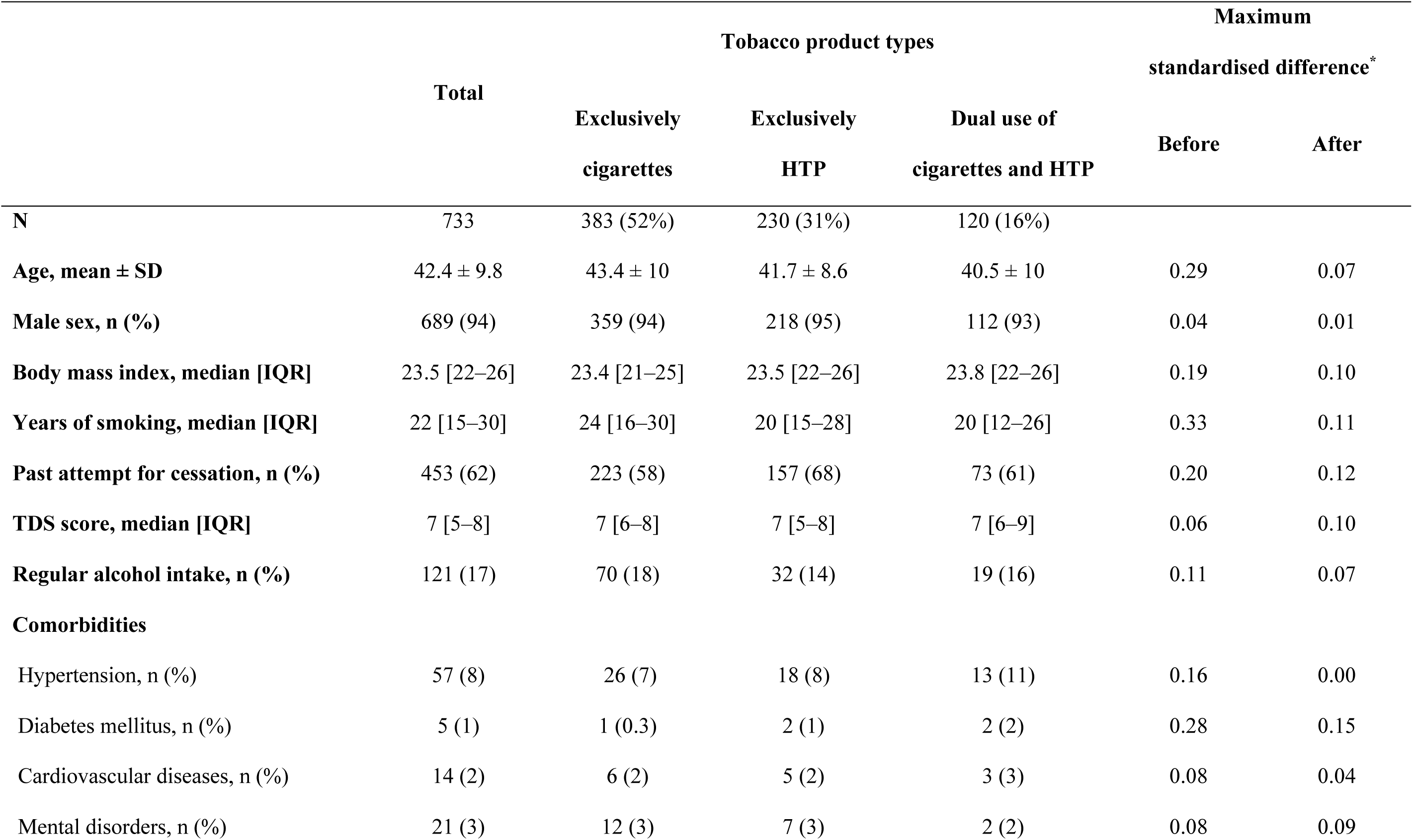
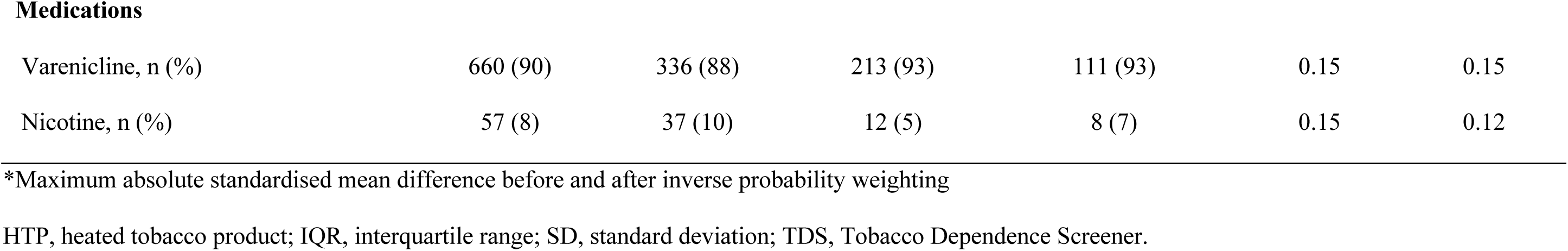
Baseline characteristics among exclusively cigarette smokers, exclusively heated tobacco product users, and dual product users.

|  | Total | Tobacco product types |  |  | Maximum standardised difference* |  |
| --- | --- | --- | --- | --- | --- | --- |
|  |  | Exclusively cigarettes | Exclusively HTP | Dual use of cigarettes and HTP | Before | After |
| **N** | 733 | 383 (52%) | 230 (31%) | 120 (16%) |  |  |
| **Age, mean ± SD** | 42.4 ± 9.8 | 43.4 ± 10 | 41.7 ± 8.6 | 40.5 ± 10 | 0.29 | 0.07 |
| **Male sex, n (%)** | 689 (94) | 359 (94) | 218 (95) | 112 (93) | 0.04 | 0.01 |
| **Body mass index, median [IQR]** | 23.5 [22–26] | 23.4 [21–25] | 23.5 [22–26] | 23.8 [22–26] | 0.19 | 0.10 |
| **Years of smoking, median [IQR]** | 22 [15–30] | 24 [16–30] | 20 [15–28] | 20 [12–26] | 0.33 | 0.11 |
| **Past attempt for cessation, n (%)** | 453 (62) | 223 (58) | 157 (68) | 73 (61) | 0.20 | 0.12 |
| **TDS score, median [IQR]** | 7 [5–8] | 7 [6–8] | 7 [5–8] | 7 [6–9] | 0.06 | 0.10 |
| **Regular alcohol intake, n (%)** | 121 (17) | 70 (18) | 32 (14) | 19 (16) | 0.11 | 0.07 |
| **Comorbidities** |  |  |  |  |  |  |
| Hypertension, n (%) | 57 (8) | 26 (7) | 18 (8) | 13 (11) | 0.16 | 0.00 |
| Diabetes mellitus, n (%) | 5 (1) | 1 (0.3) | 2 (1) | 2 (2) | 0.28 | 0.15 |
| Cardiovascular diseases, n (%) | 14 (2) | 6 (2) | 5 (2) | 3 (3) | 0.08 | 0.04 |
| Mental disorders, n (%) | 21 (3) | 12 (3) | 7 (3) | 2 (2) | 0.08 | 0.09 |
| **Medications** |  |  |  |  |  |  |
| Varenicline, n (%) | 660 (90) | 336 (88) | 213 (93) | 111 (93) | 0.15 | 0.15 |
| Nicotine, n (%) | 57 (8) | 37 (10) | 12 (5) | 8 (7) | 0.15 | 0.12 |

\*Maximum absolute standardised mean difference before and after inverse probability weighting
HTP, heated tobacco product; IQR, interquartile range; SD, standard deviation; TDS, Tobacco Dependence Screener.

### Overall success rates of the telemedicine smoking cessation programme

First, we evaluated the overall smoking cessation success rates of the telemedicine smoking cessation programme using all program participants (Figure 3). Point abstinence rates were 73.7% (95% confidence interval [CI], 70–77) at week 12 (equal to CAR9-12), 57.3% (CI, 54– 61) at week 24, 50.3% (CI, 47–54) at week 36, and 54.4% (CI, 51–58) at week 52. Additionally, CAR9-24, and CAR9-52 were 55.8% (CI, 52–59), and 42.4% (CI, 39–46), respectively.

**Figure 3.**
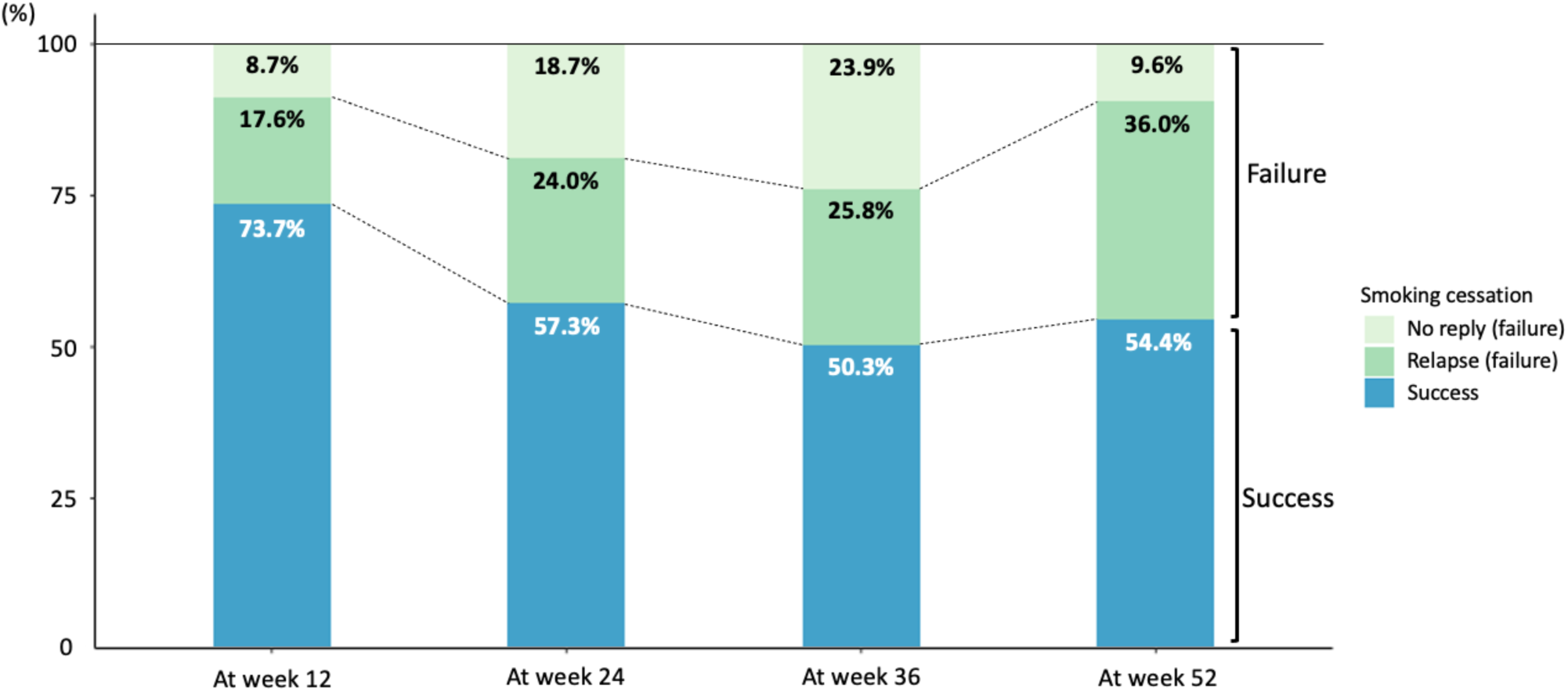
Point abstinence of telemedicine smoking cessation programme at weeks 12, 24, 36, and 52.

### Smoking cessation success rates by smoking product types

Next, we assessed the smoking cessation success rates of the telemedicine smoking cessation programme by smoking product types standardised with inverse probability weights (Table 2 and Figure 4). For continuous abstinence, the exclusively HTP group had higher rates compared with the exclusively cigarettes group in CAR9-24 (53.8% for cigarettes vs. 67.0% for HTP; adjusted odds ratio [aOR], 1.12; CI, 1.02 to 1.23; p = 0.02) and in CAR9-52 (41.0% for cigarettes vs. 50.9% for HTP; aOR 1.09; CI, 0.99 to 1.19; p = 0.08). The point abstinence rates of the HTP group at week 24 was significantly greater than those of the cigarette group. Although the differences of abstinence rates between the groups was slightly attenuated at 52 weeks, the exclusively HTP group exhibited consistent higher abstinence rates over the exclusively cigarettes group.

**Table 2.** Smoking cessation success rates of the telemedicine smoking cessation programme by smoking product types standardised with inverse probability weights.

|  | Exclusively<br>cigarettes | Exclusively<br>HTP | Adjusted OR<br>for cigarettes | P-value<br>(HTP vs Cigarettes) | Dual use of<br>Cigarettes and HTP | Adjusted OR<br>for cigarettes | P-value<br>(Dual vs Cigarettes) |
| --- | --- | --- | --- | --- | --- | --- | --- |
| **Point abstinence rates** |  |  |  |  |  |  |  |
| PA at week 12 | 70.8% [66–75] | 80.9% [76–86] | 1.08 [0.99–1.17] | 0.07 | 69.2% [61–77] | 0.96 [0.85–1.07] | 0.46 |
| PA at week 24 | 55.4% [50–60] | 67.8% [62–74] | 1.11 [1.01–1.21] | 0.03* | 43.3% [34–52] | 0.86 [0.77–0.96] | 0.008* |
| PA at week 36 | 48.3% [43–53] | 58.2% [52–65] | 1.08 [0.99–1.19] | 0.09 | 41.7% [33–50] | 0.91 [0.81–1.02] | 0.09 |
| PA at week 52 | 52.0% [47–57] | 62.2% [57–70] | 1.09 [0.996–1.20] | 0.06 | 45.0% [36–54] | 0.91 [0.81–1.02] | 0.11 |
| **Continuous abstinence rates** |  |  |  |  |  |  |  |
| CAR9-24 | 53.8% [49–59] | 67.0% [61–73] | 1.12 [1.02–1.23] | 0.02* | 40.8% [32–50] | 0.85 [0.76–0.95] | 0.004* |
| CAR9-52 | 41.0% [36–46] | 50.9% [44–57] | 1.09 [0.99–1.19] | 0.08 | 30.8% [23–39] | 0.88 [0.79–0.97] | 0.01* |

\*: P-value < 0.05. Values are reported as the rates (95% CI). P-values were calculated using an inverse probability-weighted dataset with logistic regression.
CAR, continuous abstinence rate; CI, confidence interval; HTP, heated tobacco product; OR, odds ratio; PA, point abstinence.

**Figure 4.**
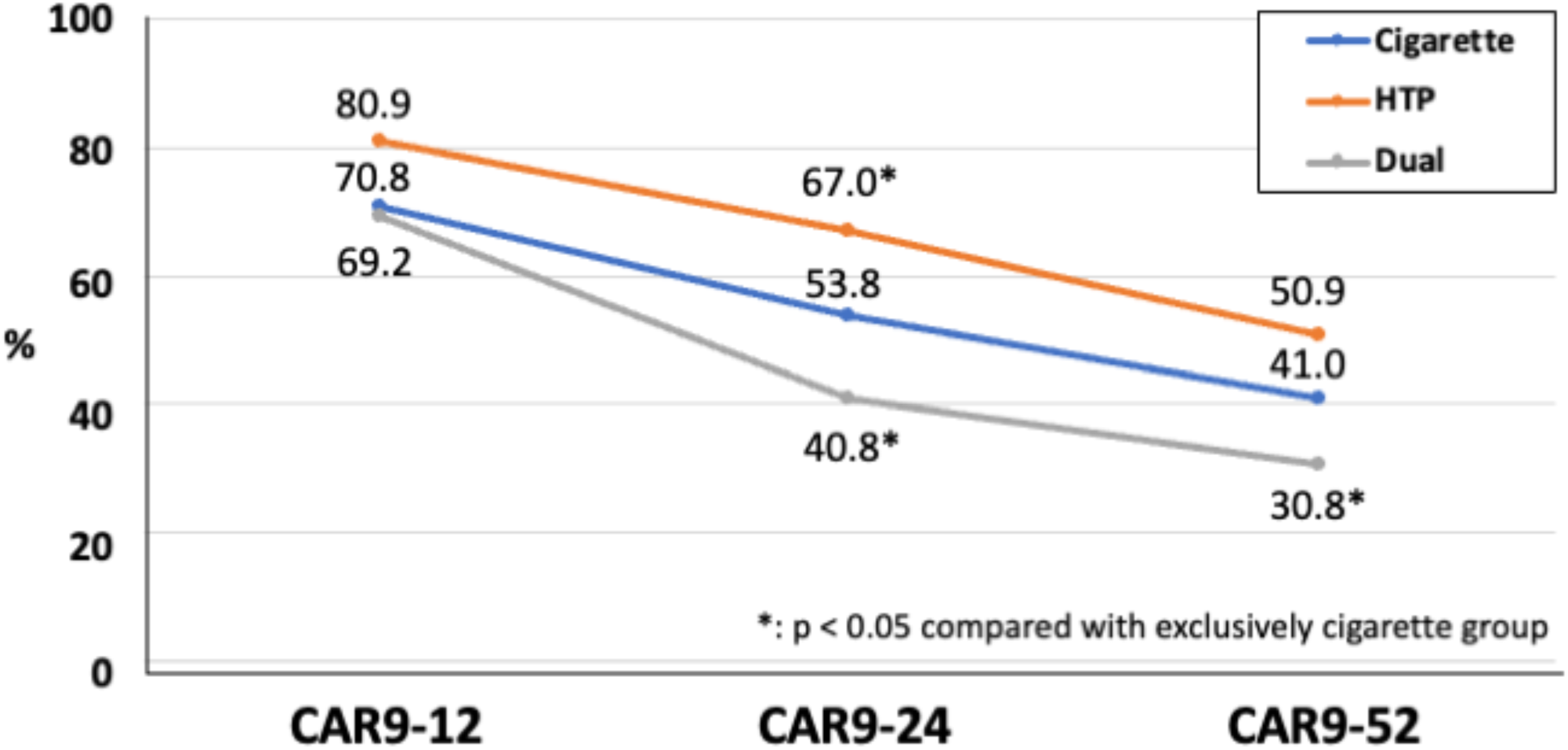
Continuous abstinence rates (CARs) from 9 to 12 weeks (CAR9-12) to 24 weeks (CAR9-24) and 52 weeks (CAR9-52) among exclusively cigarette smokers (Cigarette), exclusively HTP users (HTP), and dual users (dual).

Conversely, the dual-use of cigarettes and HTP group had lower CARs compared with the exclusively cigarettes group in both CAR9-24 (53.8% vs. 40.8% for dual use; aOR, 0.85; CI, 0.76 to 0.95; p = 0.004) and CAR9-52 (41.0% vs. 30.8% for dual use; aOR, 0.88; CI, 0.79 to 0.97; p = 0.01) (Table 2). The point abstinence rates of the dual-use group at 24 weeks were worse than those of the cigarette group.

## DISCUSSION

This study, using inverse probability weighting, assessed whether the telemedicine smoking cessation programme was effective among adult nicotine-dependent patients using HTPs. We found that: 1) overall success rates of the telemedicine smoking cessation programme were favourable in both CAR9-24 and CAR9-52; 2) exclusively HTP users had significantly higher smoking cessation success rates compared with the exclusively cigarette group in CAR9-24; and 3) dual users had significantly lower smoking cessation success rates compared with the exclusively cigarette group in CAR9-24 and CAR9-52. To our knowledge, this is the first study to elucidate the effectiveness of a telemedicine smoking cessation programme for adult smokers using HTP.

This study provides several conclusions. First, overall smoking cessation success rates by the Linkage telemedicine smoking cessation programme were favourable in both short- and long-term periods: 55.8% for CAR9-24 and 42.4% for CAR9-52. Kato et al. previously reported the effectiveness of the 6-month online smoking cessation programme in Japan for conventional cigarette smokers using online mentoring by trained smoking cessation experts, over-the-counter nicotine patches for medication, and a unique smartphone app^11^. They found that the CAR9-12 and CAR21-24 of patients who received the ascure online program were 48.6% and 47.5%, respectively, which were almost consistent with our results. Additionally, the smoking abstinence rate was around 50% after completing the 12-week Japanese smoking cessation programme through outpatient services^17,18^. Although the demographic characteristics and smoking cessation support medications were different, our telemedicine smoking cessation programme results might be reasonable compared with this data. Second, smoking product types may affect the telemedicine smoking cessation success rates, especially among exclusively HTP users who had a higher smoking cessation success rate for up to one year than the exclusively cigarettes group. Since HTPs have been promoted as a ‘switching’ product for quitting cigarette smoking by HTP manufacturers^6,19^, HTP users may have considerable motivation to quit smoking. The lower quartile of smoking years in exclusively HTP users exceeded 15 years, indicating that most had smoked cigarettes before HTP emerged (∼5 years), successfully quit cigarette smoking at some point, and switched exclusively to HTPs. Additionally, Lau et al. reported that exclusively HTP users had a lower nicotine dependence prevalent than daily exclusively cigarette smokers^7^. This characteristic of HTP users could explain our results because the severity of nicotine dependence was strongly associated with smoking cessation success ^20^.

Third, unlike exclusively HTP users, the smoking cessation success rates of the dual-use group (using both cigarettes and HTPs) were significantly lower than those of exclusively cigarette users. This result might be reasonable because dual users needed to quit both cigarette smoking and HTPs, which could be more difficult than quitting individually. Moreover, this result was consistent with the report from Ryu et al. that dual users of cigarettes and HTPs had a lower intention to quit smoking within one month than exclusively cigarette smokers^21^. Furthermore, dual users might have a different clinical profile that could attenuate smoking cessation success compared to exclusively cigarette or exclusively HTP users. Our study indicated that dual users were younger and had a shorter smoking duration than exclusively cigarette smokers. Since the age participants began smoking was one of the predictors of smoking cessation success^20^, a younger age, more susceptible to a new tobacco product, might play a critical role in recurrence. However, we did not have data for dual users who quit cigarettes or HTPs during the study period. Since the definition of abstinence for dual users was more stringent than exclusively cigarette smokers or exclusively HTP users, abstinence rates for dual users could be different if we compared continuous ‘cigarette’ abstinence rates between exclusively cigarette smokers and dual users.

The strength of this study is that it is the first study to assess the effectiveness of the telemedicine smoking cessation programme among HTP users and dual users. In the era of the coronavirus (COVID-19) pandemic, quitting smoking while minimizing person-to-person contact could be essential to reduce the risk of COVID-19 infection and severe or critical COVID-19 symptom exacerbation^22^. This study showed that the effectiveness of the telemedicine smoking cessation programme may provide an option for both cigarette and HTP users to quit smoking regardless of social contact. The limitations of this study were as follows: 1) CARs were assessed by questionnaire-based self-reported abstinence, but not biochemically validated, which could misclassify smoking status or overestimate abstinence rates; 2) since our group previously reported that more than two-thirds of HTP or e-cigarette users simultaneously smoked cigarettes^3^, some dual product users might mix in exclusively cigarette or exclusively heated tobacco groups because they were grouped by the online questionnaire, which could influence the results; 3) we excluded participants unavailable for follow-up surveys in the 12 months after, which could potentially affect adherence and success rates of the telemedicine smoking cessation programme.

In conclusion, exclusively HTP users achieved higher CARs over short- and long-term periods than exclusively combustible tobacco users through the telemedicine smoking cessation programmes. Conversely, dual users had slightly lower CARs in both short- and long-term periods, which may be partly explained by the stringent abstinence definition in this study. A telemedicine smoking cessation programme might be a reasonable option for exclusively HTP users.

## Data Availability

The data that support the study findings are available from the corresponding author upon reasonable request.

## Acknowledgements

We thank all the participants and medical staff for this study.

## Contributors

AN, TI, and TT contributed to the concept of this study. YM and TI acquired the data. AN, Tik, TF, and TT analysed the data and drafted the manuscript. All authors contributed to the interpretation of the results and critical review of the draft manuscript. All authors agree to the final version of the submitted manuscript.

## Patient consent for publication

Not required.

## Ethical approval

The study protocol was approved by the Kanazawa University institutional review board (2020-206 (3573)).

